# Meteorological Conditions and Covid-19 in Large U.S. Cities

**DOI:** 10.1101/2020.05.17.20104547

**Authors:** Hisato Takagi, Toshiki Kuno, Yujiro Yokoyama, Hiroki Ueyama, Takuya Matsushiro, Yosuke Hari, Tomo Ando

## Abstract

To determine whether prevalence of Coronavirus disease 2019 (Covid-19) is modulated by meteorological conditions, we herein conducted meta-regression of data in large U.S. cities. We selected 33 large U.S. cities with a population of >500,000. The integrated numbers of confirmed Covid-19 cases in the country to which the city belongs on 14 May 2020, the estimated population in 2019 in the country, and monthly meteorological conditions at the city for 4 months (from January to April 2020) were obtained. Meteorological conditions consisted of mean temperature (F), total precipitation (inch), mean wind speed (MPH), mean sky cover, and mean relative humidity (%). Monthly data for 4 months were averaged or integrated. The Covid-19 prevalence was defined as the integrated number of Covid-19 cases divided by the population. Random-effects meta-regression was performed by means of OpenMetaAnalyst. In a meta-regression graph, Covid-19 prevalence (plotted as the logarithm transformed prevalence on the y-axis) was depicted as a function of a given factor (plotted as a meteorological datum on the x-axis). A slope of the meta-regression line was significantly negative (coefficient, −0.069; P < 0.001) for the mean temperature and significantly positive for the mean wind speed (coefficient, 0.174; P = 0.027) and the sky cover (coefficient, 2.220; P = 0.023). In conclusion, lower temperature and higher wind speed/sky cover may be associated with higher Covid-19 prevalence, which should be confirmed by further epidemiological researches adjusting for various risk and protective factors (in addition to meteorological conditions) of Covid-19.

## Introduction

Higher temperature and ultraviolet (UV) index in Northern Europe have been reported as the most important meteorological protective factors for the transmission of influenza virus.^1^ On the other hand, a recent study in China suggests that higher temperature and UV radiation may not be associated with a decrease in the epidemics of Coronavirus disease 2019 (Covid-19).^2^ To determine whether prevalence of COVID-19 is modulated by meteorological conditions, we herein conducted meta-regression of data in large U.S. cities.

## Methods

We selected 33 large U.S. cities with a population of >500,000 in 2010 from the U.S. Census Bureau (http://www.census.gov). The integrated numbers of confirmed Covid-19 cases in the country to which the city belongs on 14 May 2020 from the Johns Hopkins Coronavirus Resource Center (https://coronavirus.jhu.edu), the estimated population in 2019 in the country from the U.S. Census Bureau, and monthly meteorological conditions at the city for 4 months (from January to April 2020) from the National Weather Service (https://www.weather.gov) were obtained (Table 1). Meteorological conditions consisted of mean temperature (F), total precipitation (inch), mean wind speed (MPH), mean sky cover, and mean relative humidity (%). Monthly data for 4 months were averaged or integrated. The Covid-19 prevalence was defined as the integrated number of Covid-19 cases divided by the population. Random-effects meta-regression was performed by means of OpenMetaAnalyst (http://www.cebm.brown.edu/openmeta/index.html). In a meta-regression graph, Covid-19 prevalence (plotted as the logarithm transformed prevalence on the y-axis) was depicted as a function of a given factor (plotted as the meteorological data on the x-axis).

**Table 1.** Extracted data in each large U.S. city and country to which the city belongs.

| City | State | County | Covid-19 prevalence in the country |  |  | Meteorological condition at the city |  |  |  |  |
| --- | --- | --- | --- | --- | --- | --- | --- | --- | --- | --- |
|  |  |  | Cases (N) | Population (N) | Prevalence | Mean temperature (F) | Total precipitation (inch) | Mean wind speed (MPH) | Mean sky cover | Mean relative humidity (%) |
| Albuquerque | New Mexico | Bernalillo | 1,149 | 679,121 | 0.00169 | 47.1 | 1.65 | 8.4 | 0.45 | 43 |
| Austin | Texas | Travis | 2,345 | 1,273,954 | 0.00184 | 60.6 | 12.06 | 8.2 | 0.63 | 72 |
| Baltimore | Maryland | Baltimore | 4,290 | 827,370 | 0.00519 | 46.5 | 14.66 | 7.4 | 0.67 | 61 |
| Boston | Massachusetts | Suffolk | 15,881 | 803,907 | 0.01975 | 40.6 | 12.62 | 12.1 | 0.62 | 58 |
| Charlotte | North Carolina | Mecklenburg | 2,342 | 1,110,356 | 0.00211 | 53.7 | 21.33 | 7.3 | 0.62 | 63 |
| Chicago | Illinois | Cook | 58,457 | 5,150,233 | 0.01135 | 37.9 | 10.86 | 10.3 | – | – |
| Columbus | Ohio | Franklin | 4,227 | 1,316,756 | 0.00321 | 41.6 | 19.31 | 8.9 | 0.75 | 67 |
| Dallas | Texas | Dallas | 6,837 | 2,635,516 | 0.00259 | 57.0 | 17.53 | 11.1 | 0.65 | 70 |
| Denver | Colorado | Denver | 4,359 | 727,211 | 0.00599 | 38.0 | 2.82 | 10.1 | 0.57 | 57 |
| Detroit | Michigan | Wayne | 18,770 | 1,749,343 | 0.01073 | 37.5 | 11.35 | 9.6 | 0.81 | 73 |
| D.C. | D.C. | – | 6,736 | 705,749 | 0.00954 | 48.7 | 14.61 | 9.1 | 0.70 | 63 |
| El Paso | Texas | El Paso | 1,456 | 839,238 | 0.00173 | 57.1 | 3.07 | 8.7 | 0.45 | 40 |
| Fort Worth | Texas | Tarrant | 4,076 | 2,102,515 | 0.00194 | 57.0 | 17.53 | 11.1 | 0.65 | 70 |
| Houston | Texas | Harris | 8,817 | 4,713,325 | 0.00187 | 64.2 | 13.90 | 8.5 | 0.68 | 71 |
| Indianapolis | Indiana | Marion | 7,793 | 964,582 | 0.00808 | 41.4 | 15.61 | 10.5 | 0.74 | 73 |
| Jacksonville | Florida | Duval | 1,215 | 957,755 | 0.00127 | 65.2 | 6.36 | 7.4 | 0.60 | 68 |
| Las Vegas | Nevada | Clark | 5,045 | 2,266,715 | 0.00223 | 58.1 | 2.31 | 6.5 | 0.45 | 37 |
| Los Angeles | California | Los Angeles | 35,392 | 10,039,107 | 0.00353 | 61.0 | 7.17 | 7.1 | 0.48 | 62 |
| Louisville | Kentucky | Jefferson | 1,741 | 766,757 | 0.00227 | 47.9 | 16.28 | 8.6 | 0.75 | 62 |
| Memphis | Tennessee | Shelby | 3,542 | 937,166 | 0.00378 | 52.9 | 27.76 | 8.8 | 0.68 | 71 |
| Milwaukee | Wisconsin | Milwaukee | 4,387 | 945,726 | 0.00464 | 35.4 | 10.52 | 10.4 | 0.68 | 66 |
| Nashville | Tennessee | Davidson | 3,745 | 694,144 | 0.00540 | 51.0 | 23.11 | 7.8 | 0.70 | 64 |
| New York City | New York | New York City | 188,545 | 8,336,817 | 0.02262 | 44.4 | 12.74 | 6.5 | – | 57 |
| Oklahoma City | Oklahoma | Oklahoma | 1,013 | 797,434 | 0.00127 | 49.3 | 10.86 | 12.1 | – | – |
| Philadelphia | Pennsylvania | Philadelphia | 19,093 | 1,584,064 | 0.01205 | 45.1 | 12.79 | 9.8 | 0.67 | 60 |
| Phoenix | Arizona | Maricopa | 6,599 | 4,485,414 | 0.00147 | 63.6 | 3.55 | 5.9 | – | 42 |
| Portland | Oregon | Multnomah | 940 | 812,855 | 0.00116 | 47.7 | 12.35 | 7.2 | 0.68 | 71 |
| San Antonio | Texas | Bexar | 1,976 | 2,003,554 | 0.00099 | 63.0 | 7.35 | 8.3 | 0.65 | 66 |
| San Diego | California | San Diego | 5,391 | 3,338,330 | 0.00161 | 61.0 | 6.69 | 5.1 | 0.55 | 69 |
| San Francisco | California | San Francisco | 1,999 | 881,549 | 0.00227 | 55.0 | 3.86 | 9.3 | 0.54 | 69 |
| San José | California | Santa Clara | 2,391 | 1,927,852 | 0.00124 | 55.2 | 4.06 | 5.8 | 0.51 | 65 |
| Seattle | Washington | King | 7,290 | 2,252,782 | 0.00324 | 46.4 | 18.15 | 9.0 | 0.73 | 73 |
| Tucson | Arizona | Pima | 1,696 | 1,047,279 | 0.00162 | 59.7 | 2.09 | 6.8 | 0.11 | 40 |

## Results

Results of the meta-regression were summarized in **Table 2**. A slope of the metaregression line was significantly negative (coefficient, –0.069; P < 0.001; **Figure 1**, upper panel) for the mean temperature (indicating that Covid-19 prevalence decreased significantly as the mean temperature increased) and significantly positive for the mean wind speed (coefficient, 0.174; P = 0.027; **Figure 1**, middle panel) and the sky cover (coefficient, 2.220; P = 0.023; **Figure 1**, lower panel) (indicating that Covid-19 prevalence increased significantly as the mean wind speed and the sky cover increased).

**Figure 1.**
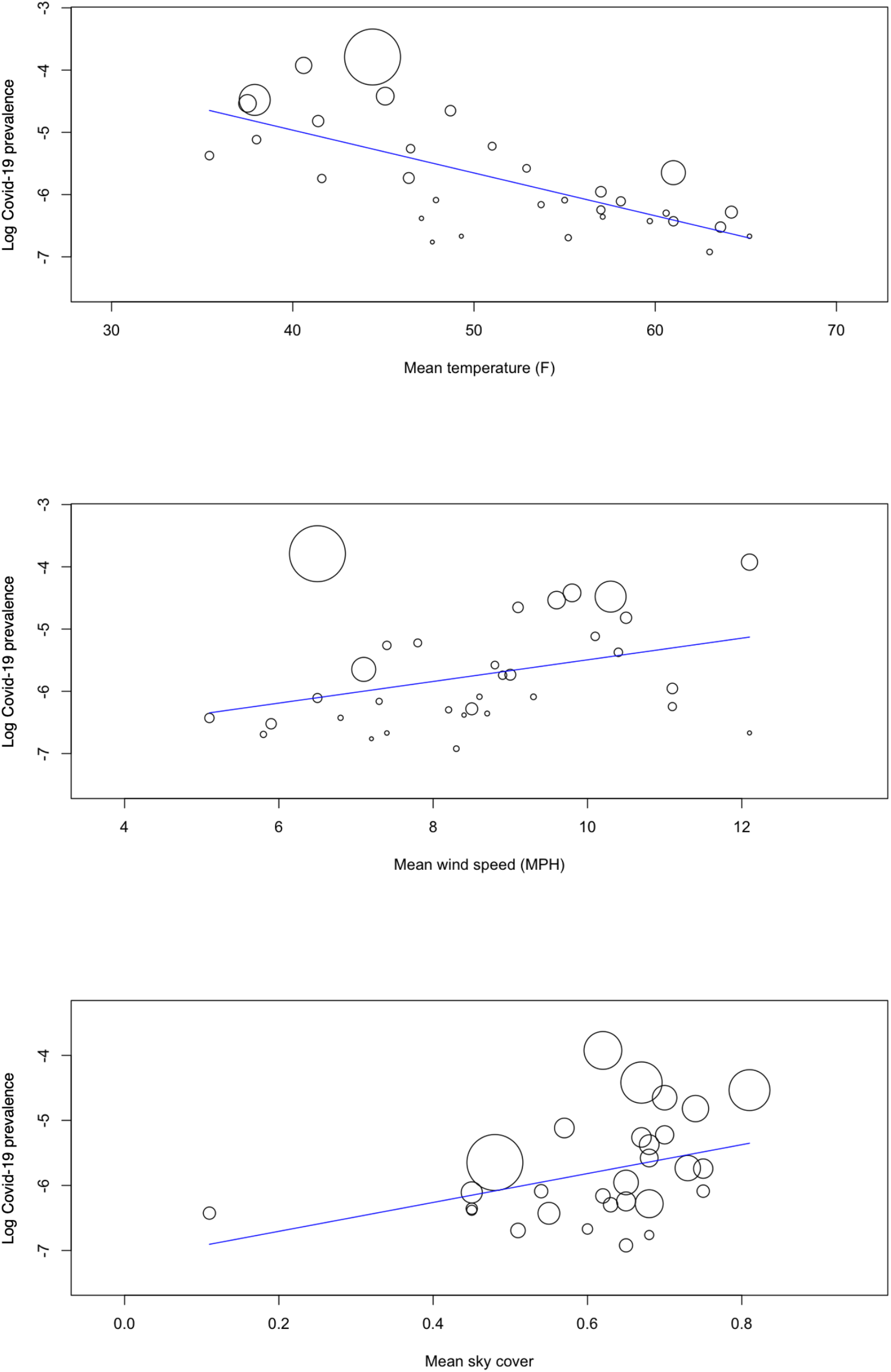
Meta-regression graph depicting covid-19 prevalence (plotted as the logarithm transformed prevalence on the y-axis) as a function of the given factors (plotted as the meteorological data on the x-axis).

## Discussion and Conclusion

The present meta-regression suggests that temperature may be negatively and wind speed/sky cover may be positively associated with COVID-19 prevalence. Higher sky cover is probably related to lower UV radiation. Our recent preliminary study of Japanese prefectural data suggests that temperature and UV index may be negatively associated with COVID-19 prevalence,^3^ which could strengthen the present findings. Despite the association of lower temperature and UV index with the influenza transmission,^1^ no association of temperature and UV radiation with the COVID-19 epidemics has been reported,^2^ however, which may be denied by the present results of the association of lower temperature and higher sky cover with higher COVID-19 prevalence. In conclusion, lower temperature and higher wind speed/sky cover may be associated with higher Covid-19 prevalence, which should be confirmed by further epidemiological researches adjusting for various risk and protective factors (in addition to meteorological conditions) of Covid-19.

## Data Availability

The datasets generated during and/or analysed during the current study are available from the corresponding author on reasonable request.

---

**Supplementary Table 2.** Meta-regression summary.

| Covariate | Coefficient |  |  | Standard error | P value |
| --- | --- | --- | --- | --- | --- |
|  |  | Lower bound | Upper bound |  |  |
| Mean temperature (F) | −0.069 | −0.093 | −0.045 | 0.012 | <0.001 |
| Total precipitation (inch) | 0.038 | −0.004 | −0.081 | 0.022 | 0.075 |
| Mean wind speed (MPH) | 0.174 | 0.020 | 0.328 | 0.079 | 0.027 |
| Mean sky cover | 2.220 | 0.313 | 4.128 | 0.973 | 0.023 |
| Mean relative humidity (%) | 0.007 | −0.020 | 0.035 | 0.014 | 0.613 |

